# Comparative Immune profiling in Pancreatic Ductal Adenocarcinoma Progression Among South African patients

**DOI:** 10.1101/2023.10.23.23297385

**Authors:** Nnenna Elebo, Ebtesam A. Abdel-Shafy, Jones A.O. Omoshoro-Jones, Zanele Nsingwane, Ahmed A.A. Hussein, Martin Smith, Geoffrey Candy, Stefano Cacciatore, Pascaline Fru, Ekene Emmanuel Nweke

## Abstract

**Introduction:** Pancreatic Ductal Adenocarcinoma (PDAC) is an aggressive cancer with a 5-year survival rate of only 11%. PDAC is characterized by an immunosuppressive microenvironment; thus, there have been multiple attempts to target it, although with little success. A better understanding of the immune landscape in PDAC is required to help elucidate the roles of these cells for effective targeting. This study investigated the expression of circulating key immune cell markers in South African PDAC patients.

**Method:** Blood samples were obtained from a total of 34 PDAC patients consisting of 22 resectable (RPC), 8 locally advanced (LAPC) and 4 metastatic (MPC), 6 Chronic Pancreatitis (CP), and 6 healthy volunteers (HC). Immunophenotyping, real-time polymerase chain reactions (PCR), enzyme-linked immunosorbent assay (Elisa), and reactive oxygen species (ROS) assays were conducted. Statistical analysis was conducted in R (version 3.6.1) and Wilcoxon and Kruskal-Wallis rank-sum tests were used to compare between groups. Kaplan-Meier analysis and Spearman’s rank test were used for survival and correlation analyses, respectively.

**Results:** Granulocyte and neutrophil levels were significantly elevated while lymphocytes decreased with PDAC severity. The total percentages of CD4^+^, CD8^+^, and CD3^+^CD4^-^CD8^-^ T-cells increased across the group. Of note are the reduction of CD16^+^ NKTs across the RPC (*p* = 0.002), LAPC (*p* = 0.01), and MPC (*p* = 0.017) groups when compared to HC. Both NK (*p* = 0.0047) and NKTs (*p* = 0.0027) increased in RPC but decreased in both LAPC and MPC when compared to HC. Although there was no statistical correlation or differences observed when comparing the PDAC groups with the control groups, RPC had the highest foldchange for both *CD4* (11.75 ± 44.31) and *CD3* (30.47 ± 75.01) while the LAPC group had the highest fold change for *CD8* (3.86 ± 7.35) and *CD16* (51.69 ± 108.9) genes compared to MPC. The inflammatory status of PDAC was assessed by DEPPD levels of serum which were elevated in RPC (*p* = 0.003) and LAPC (*p* = 0.008) but decreased in MPC (*p* = 0.025), compared to the HC group. ROS was shown to be positively correlated with GlycA (R=0.45, *p* = 0.00096).

**Conclusion:** The expression of these immune cell markers observed in this pilot study provides insight into their potential roles in tumour progression in the patient group and suggests their potential utility in the development of immunotherapeutic strategies.

## INTRODUCTION

PDAC is the most common neoplasm of the pancreas with a very poor prognosis and survival rate of <12% in 5 years (1). PDAC has been projected to be the second leading cause of cancer worldwide (2). Due to the late presentation, most patients are diagnosed with a locally advanced or metastatic stage of the disease which precludes the chance of surgical resection (3). Treatment strategies include chemotherapy, radiotherapy, immunotherapy and surgery (4,5). Currently, surgery is the best clinical treatment for PDAC, however, only about 15-20% of patients will undergo surgery leaving a majority with limited options (6). The role of the immune system and its interaction with PDAC cells has emerged as a pivotal focus of the investigation. Immune cells have dual roles in PDAC which are contributing to the tumour progression and paradoxically offering avenues for therapeutic intervention. The tumour microenvironment (TME) of PDAC is characterised by malignant cells, stromal components, and immune cells which converge in a delicate balance (7,8). Neutrophils play diverse roles in pancreatic cancer such as angiogenesis, progression, metastasis, and immunosuppression (9). Blood neutrophil to lymphocyte ratio (NLR) has been associated with prognosis in pancreatic cancer (10). Effector immune cells such as natural killer cells, CD4^+^ T-cells, and CD8^+^ T-cells exert intricate influences on the tumour behaviour either by fueling its growth or orchestrating its suppression (11). Immunosuppressive mechanisms of PDAC cells include editing the immune system to become unrecognizable leading to tumour escape, activation, and release of immunosuppressive molecules such as IL-10 and TGFβ which inhibit immune response and promote tumour growth and metastasis (12). Pancreatic cancer cells downregulate the expression of MHC class I molecules by interfering with the antigen cross-presentation to effector T-cells, further exacerbating cancer (13).

Effector immune cells are vital because they are present and activated at the early stages of PDAC. The specificity of CD3 antigen for T cells and its appearance at all stages of T cell development makes it an ideal T cell marker for both detection of normal T cells and T cell neoplasms. T-helper cells (CD4^+^) and T-cytotoxic cells (CD8^+^ T cells) form large proportions of the CD3^+^T cells involved in cell-mediated immunity (14). Alteration of either the number or the function of CD4^+^ T cells and CD8^+^ T cells will affect the immune response. Hence maintaining the balance between CD4^+^ and CD8^+^ T cells is critical for tumour immunity. Cytotoxic lymphocytes play important roles in innate and adaptive immune system response against tumour by secreting cytokines to facilitate their anti-tumour effect (15). NK cells represent about 5-25% of circulating lymphocytes and express CD16, CD56, and CD57 markers in humans (16). The NK populations are distinguished by the markers CD56brightCD16^-^ for immune modulatory function via the interferon-γ (IFN-γ) secretion and CD56dimCD16^+^ for cytotoxic abilities (17).

Immuno-inflammatory response plays a vital role in tumour growth and progression evident from the immune dysfunction as observed in PDAC (18,19). This systematic inflammatory response could be quantified through different scores such as ratios between different circulating immune cells (20) and correlation between immune cells and reactive oxygen species (ROS). Additionally, immune cells induce ROS production through the secretion of tumour necrosis factor-α (TNF-α). ROS have been shown to exert an immunosuppressive effect on NK and T-cells (21). PDAC accumulates ROS which has dual roles depending on their concentration (22,23). It facilitates cancer progression at mild to moderate levels (24) while excessive ROS production promotes the release of cytochrome c into the cytoplasm which mediates programmed cell death (25). Immuno-inflammatory responses have been shown to differ between population groups because of ancestral and environmental factors (26). Furthermore, a recent study from our laboratory showed that the inflammatory markers GlycA and GlycB were significantly elevated in PDAC patients of African descent when compared to healthy individuals (27). These differential immune responses contribute to cancer disparities because of their impacts on cancer progression (28). In this study, we revealed the immune response at both mRNA and protein levels in different stages of PDAC patients of African ancestry.

## METHODS

### Patient Recruitment

Ethics clearance for this study was obtained from the Human Research Ethics Committee of the University of the Witwatersrand (Study number: M190681). Participants gave written informed consent and were recruited from the Hepatopancreatobiliary Unit at Chris Hani Baragwanath Academic Hospital, Soweto Johannesburg, South Africa, and sample processing was done at the Department of Surgery, Faculty of Health Science, University of the Witwatersrand. Inclusion criteria included patients 18 years old and above, of African ancestry, and diagnosed with one of the three stages of PDAC notably resectable pancreatic cancer (RPC), locally advanced pancreatic cancer (LAPC) and metastatic pancreatic cancer (MPC) and chronic pancreatitis (CP) as well as healthy volunteers for the control group. All patients self reported as of African ancestry. Patients with organ failure or undergoing immunotherapy at the time of study were excluded.

### Sampling and Processing

Fasting blood samples were collected by venepuncture in two separate clear vacutainer tubes (BD Biosciences, Franklin Lakes, NJ, USA) with coagulant EDTA (purple lid) and without anti-coagulant (red lid). The blood was processed to obtain serum by centrifuging at 1734 xg, 4 °C for 10 min after allowing it to clot for 30–60 min at room temperature. Following gravity separation for 30-60 min at room temperature, the plasma component was then isolated. For immunophenotyping assays, a subset of twenty-seven samples including 10 RPC, 6 LAPC, 4 MPC, 2 CP, and 5 healthy controls (HC) was used. White blood cells were fixed by adding 1 ml of diluted BD FACS Lyse (BD Biosciences, New Jersey, United States) solution to 100µl of the whole blood sample from the purple capped vacutainer tube within 2 h of collection. The whole blood FACS lyse mix was allowed to stand at room temperature for 12-15 mins after which it was stored at -80°C until analysis. Peripheral blood mononuclear cells (PBMCs) which comprises the major immune cells such as lymphocytes, monocytes, and macrophages were separated using Ficoll-Paque™ (GE Healthcare, Illinois, United States) separation method (29). Serum and plasma samples were processed within 2 h of collection while PBMCs at a concentration of 1×105 to 106 1844cells/ml were separated and stored in a freezing medium (10% dimethyl sulphoxide, Sigma Aldrich, Missouri, USA and 90% Gibco Bovine Serum (FBS), Thermo Fischer, Massachusetts, USA) and aliquoted (200 µl) in single-use vials, which were stored at -80 °C until needed. Samples were only thawed once to preserve integrity.

### Characterization of different immune cells using 6-colour panel flow cytometry

Multicolour flow cytometry immunophenotyping analysis was used to determine immune cell populations and frequency. A 6-colour panel was established to characterize heterogeneous cell populations in the PDAC stages and compared with the controls (**Table S1**). Fully stained samples and unstained samples were prepared from the thawed cells. Stained samples were prepared by adding antibodies in the dark at previously titrated volume and then incubated for 30 mins at room temperature. Antibodies were optimized by titration to optimally stain lymphocyte populations and their subpopulations using CD3 BD Horizon Brilliant™

Ultraviolet (BUV), CD4 Alexa flour (AF-700) and CD8 Brilliant Violet™ 605 (BV-605), CD56 PE Phycoerythrin Cyanine 7 (PECy7), CD57 (BB515) while the granulocyte population was stained with CD16 PECy5. All antibodies were obtained from BD LSRFortessa^TM^ II flow cytometer BD Biosciences, (New Jersey, United States). Instrument controls for voltage optimisation using single stained and unstained cells as well cytometer setup and tracking beads assays were performed with each experiment. Compensation controls using compensation beads (Anti-mouse Ig, _K_/Negative control compensation particles set; BD Biosciences, New Jersey, United States) to exclude spillover were also included in addition to the experimental controls of unstained samples (30). A total of 100,000 events were recorded on the flow cytometer (BD Biosciences, New Jersey, United States).

### Gene expression analysis of immune-related markers

Total RNA was extracted from PBMCs isolated from the PDAC and control samples, using the TriReagent^®^ (Sigma Aldrich, Missouri, United States) according to the manufacturer’s instructions. The quality of RNA was measured using a NanoDrop ND-1000 Spectrophotometer (Thermo Fischer Scientific, Massachusetts, United States), and A260/280 ratio > 1.8 was observed across all samples. Complimentary DNA (cDNA) synthesis was performed from 250 ng/µl of total RNA using the Photoscript^®^ II First Strand cDNA synthesis Kit E6560S (New England BioLabs^®^ Inc.), according to the manufacturer’s instructions.

A quantitative Real-time Polymerase Chain Reaction (RT-PCR) was then carried out using the TaqMan^®^ Fast Advanced Master Mix (Thermo Fischer, Massachusetts, United States) as per the manufacturers’ instructions. The reference gene Microsomal Ribosomal Protein L19 (*MRPL19*) and target genes *CD8A*, *CD4*, *CD3*, *CD16/FCGRB*, *CD56/NCAM1,* and *CD57/B3GAT* were obtained from Thermo Fischer Scientific, Massachusetts, United States. MRPL19 is a housekeeping gene used as a control because its expression does not change in pancreatic cancer (31). The MIQE guidelines were strictly adhered to (32). The Quant Studio™ 1 Real-Time System (Thermo Fischer Scientific, Massachusetts, United States) was used to run the RT-PCR reactions.

### Measurement of plasma levels of CD4 and CD8 cellular markers

An immunoassay ELISA kit which have been pre-coated with antibodies specific to human CD4 and CD8 wereused to quantify the concentration of these immune cell markers in the plasma of PDAC samples. Elisa kits were obtained from Elabscience Biotechnology Inc (Houston, USA). The tests were performed according to the manufacturer’s protocol. Standards and samples were assayed in duplicates and the optical density was determined at 430nm. The concentrations of CD4 and CD8 (ng/ml) markers were calculated from the standard curve.

### Reactive oxygen species (ROS) assessment using N, N-diethyl-para-phenylenediamine

N, N-diethyl -para-phenylenediamine (DEPPD) sulfate is a compound that reacts with the serum to form a coloured cation radical (33). The amount of radical cation formed is related to the oxidative status of serum and can be expresses as hydrogen peroxides equivalents (34).

Hydroperoxides decompose into alkoxy and peroxyl radicals, which convert N alkylated p-phenylenediamines to form coloured dye complexes depending on the redox potential of the peroxide (33). This can be measured spectrophotometrically with absorbance being proportional to the number of hydroperoxyl compounds and thus the oxidative status of the sample can be determined.

One hundred and forty microlitres of 0.1M sodium acetate buffer (pH 4.8) was added to each allocated well of a 96-well plate. Five microlitres of serum samples consisting of PDAC and controls as well as standards of different concentrations of hydrogen peroxide solutions: 50, 25, 6.25, 3.13, 1.56, 0.78, and 0.39 µM were added in duplicates. DEPPD and Iron sulfate were dissolved in 0.1M Sodium acetate buffer pH 4.8, respectively. One hundred microlitre of the reagent mixture prepared at a ratio of 1:25 was then added to each well. The solution was incubated at 37 °C for 1 min. Colour development was recorded at 505 nm at 25 °C, every 15 secs for 30 repeats using an FL 600 Microplate reader Multiscan Sky Microplate Spectrophotometer (Thermo Fisher Scientific, Massachusetts USA).

## Data Analysis

The immunophenotyping data were analysed using FlowJo LLC version 10.8 (BD, Biosciences, New Jersey, United States) using flow cytometry standard (FCS) files linked to the compensation controls from FACSDiva™ software. Cells were gated as singlets to exclude doublets using forward scatter height (FSC-H) and forward side scatter area (FSC-A) parameters. The gating strategies were optimised to identify distinct cell populations based on scatter parameters such as white blood cells into lymphocytes and granulocytes based on forward side scatter (FSC) versus side scatter as well as fluorochrome intensities conjugated to each antibody used. Subsequently, fluorescence histograms and dot plots were generated to visualize the distribution of marker expression within the defined gates. Statistical metrics, including percentages of the total parent, were computed for specific markers.

The 2^-ΔΔCT^ method was used to calculate relative changes in gene expression using the Microsoft Excel^®^ software. The RT-PCR data were analysed using GraphPad Prism™ software version 8 (GraphPad Software Inc, California, United States). A Shapiro-Wilk test was used to test for normality. Statistical analysis and graphical illustrations of the data were generated in R (version 3.6.1) and R studio (version 1.1.456) software using KODAMA. The data was non-parametric hence Wilcoxon and Kruskal–Walli’s rank-sum test was used to compare differences between the controls and PDAC groups. The ELISA data was analysed using Microsoft Excel by deducing the concentrations of the CD4 and CD8 T cells secreted in plasma from a standard curve. The R studio V1.3 software was used for ROS statistical analysis. A standard curve of the standards was plotted to calculate the concentration of the samples. Sample ROS concentrations were calculated from the standard curve and differences (p<0.05) between the PDAC samples and the combined healthy and CP groups was determined using an unpaired non-parametric Wilcoxon test. Spearman’s rank test was then used to calculate the correlation coefficient (rho) betweenthe oxidative status, immunophenotyping, and real-time PCR assays.

## RESULTS

Forty patients including 22 RPC, 8 LAPC, 4 MPC, and 6 CP as well as six age-matched healthy controls (HC) were recruited in this study. All the healthy participants confirmed that they were in good health and were not taking any regular medication, to be eligible for the study. An overview of the total number of patients recruited is shown in **Figure S1.** The clinical parameters, demographics, and comorbidities have been reported in a previous study (27).

### Effector Immune Cells Response in PDAC

To identify effective immune cell response patterns in PDAC, the immune cell markers CD3, CD4, CD8, CD16, CD56, and CD57 were assessed in the lymphocytes and granulocyte cell populations. CD3, CD4, and CD8 immune cell markers were used to target T-cell lymphocyte subpopulations, CD16 and CD56 were used to determine the NK cell levels, and CD57 for NK cell differentiation (35). Cells were gated using forward versus side angle light scatter to identify lymphocytes and granulocytes with side scatter versus the various stained markers to confirm these populations as shown in **Figure 1**. The lymphocytes population was further used to identify CD3^+^CD8^+^ (cytotoxic), CD3^+^CD4^+^ (helper), NKs, NKTs, and CD3^+^CD4^-^ CD8^-^ (Double Negative) T cells while the granulocytes were used to identify CD16^+^ neutrophil subpopulations (**Figure 1**). The gating strategy used also showed the other subpopulations such as CD16^+^NKs and CD57^+^NKs derived from NK subsets and CD8^+^CD57^+^ derived from cytotoxic T-cells.

**Figure 1:**
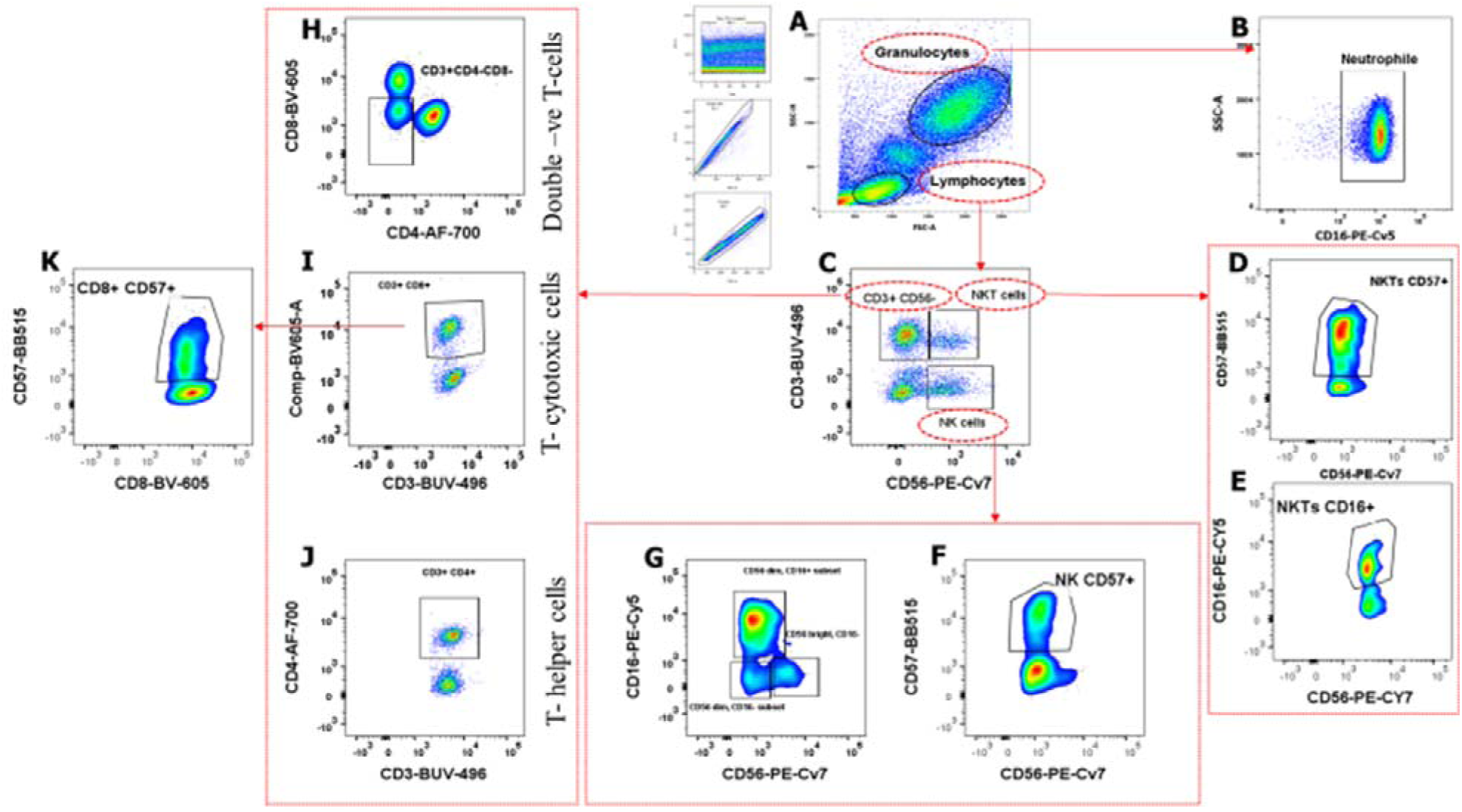
Gating strategy of the immune cell markers populations and subsets using FlowJo. **A.** The total number of cell events collected was 100,000. Cells were gated into Singlets (FSC-A versus FSC-H) which was further gated to granulocyte and lymphocyte populations. **B.** Granulocytes is further gated to Neutrophils (SSC-A versus CD16). **C.** Lymphocytes were gated to CD3CD56^+^, NKT-cells and NK cells (CD3 versus CD56) **D and E**. NKT cells is further gated into CD16^+^NKTs subsets (CD57 versus CD56) and CD57^+^NKTs subsets (CD16 versus CD56) respectively. **F and G.** NK cells further gated to CD57^+^ NK subsets and CD56^+^CD16 NK subsets **H.I.and J.** CD3^+^ T-cells was gated to different subsets; double negative T-cells (CD3^+^CD4^-^CD8^-^), T-cytotoxic cells and T-helper cells. **K**.T-cytotoxic cells was gated into CD8^+^CD57^+^ (CD57 versus CD8). CD3 BD Horizon Brilliant™ Ultraviolet (BUV), CD4 Alexa flour (AF-700) and CD8 Brilliant Violet™ 605 (BV-605), CD56 PE Phycoerythrin Cyanine 7 (PECy7), CD57 (BB515) while the granulocyte population was stained with CD16 PECy5

The total percentages of granulocytes and neutrophils significantly increased while CD4^+^, CD8^+^, and CD3^+^CD4^-^CD8^-^ T-cells decreased across the group (**Figure 2**). Additionally, CD4^+^ and CD8^+^T-cells levels changed across the groups, compared to HC. The HC group had the highest percentage of lymphocytes and CD3^+^CD4^+^ T-cell subset. It should be noted note that the downregulated CD4^+^ T-cell level in this cohort is not related to HIV status because the CD4 counts of these patients were above 300 cells/µl which is within the normal range. These patients are on antiretroviral (ARV) treatments and have normal CD4 counts as shown previously (27). Conversely, the LAPC group has the highest concentration of CD3^+^CD8^+^ T-helper cells (**Table S2**). Additionally, NKT CD16^+^ levels decreased across the PDAC groups when compared to both CP and HC. Although not significant both NKs and NKTs were reduced in LAPC and MPC when compared to the controls (**Figure 2**).

**Figure 2:**
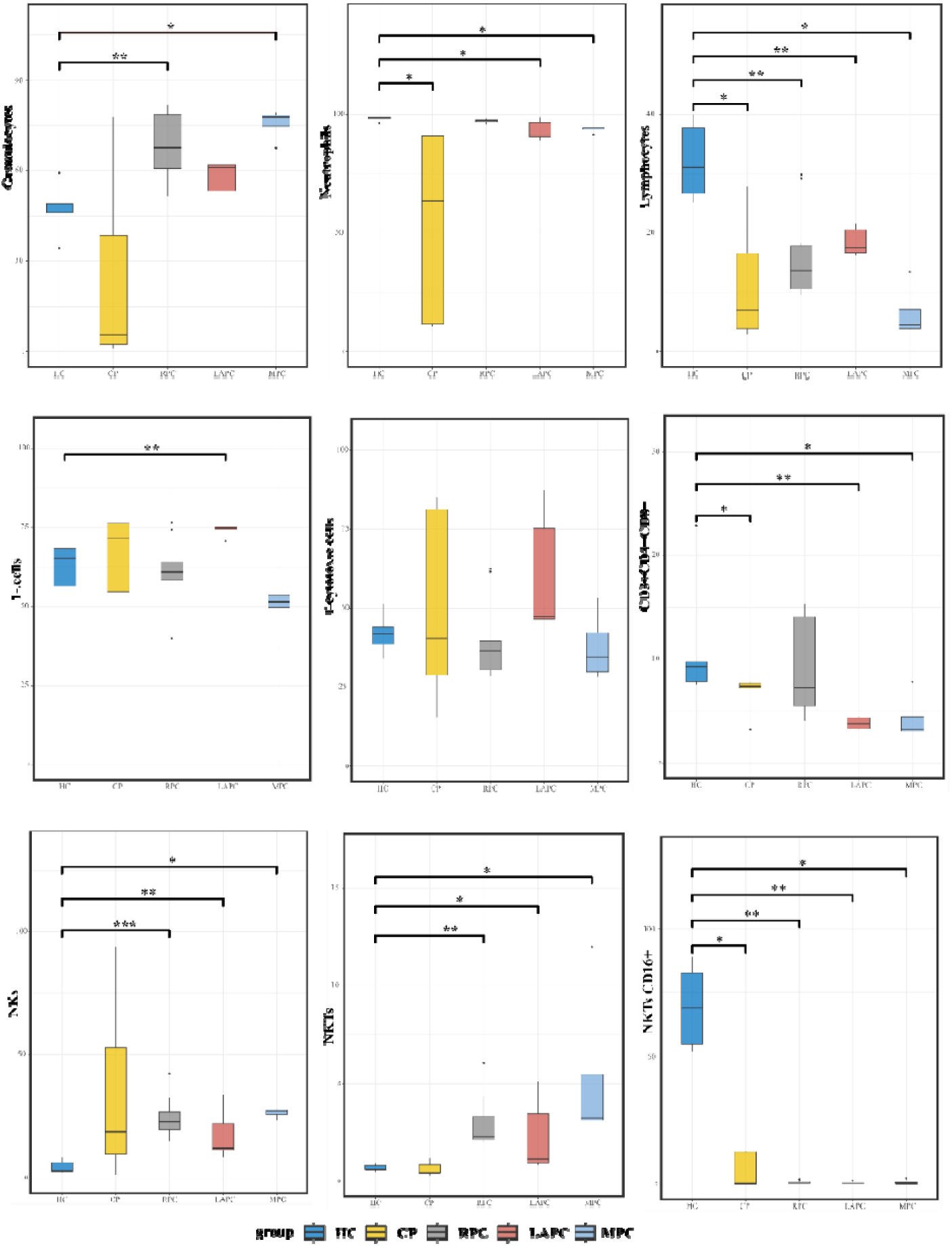
Boxplots showing the comparison of the total percentage population of the immune cell markers across the PDAC and control groups. The total percentage of granulocytes, lymphocytes, and neutrophils populations was significantly altered across all the PDAC groups, while T-cells, T-helper cells, and double negative T-cells (CD3^+^CD4^-^CD8^-^) were significantly changed in RPC and MPC when compared to HC. Furthermore, the total percentage of both NKs and NKTs was significantly increased in RPC when compared to the HC. CD: Cluster of differentiation., HC: Healthy Controls; CP: Chronic Pancreatitis; RPC: Resectable Pancreatic Ductal Adenocarcinoma; LAPC: Locally Advanced Pancreatic Ductal Adenocarcinoma; MPC: Metastatic Pancreatic Ductal Adenocarcinoma. *p < 0.05, **p < 0.01, ***p < 0.001, n.s., not significant

To understand the immune cell response across the PDAC groups, a heatmap was used to compare their frequencies (Figure 3A). The HC and CP groups were shown to have the highest intensities of the effective immune cells while the PDAC groups had the lowest. Furthermore, to further understand the relationship between the immune cells and PDAC progression, the correlation matrix was used to assess the intra-correlation of these immune cells (Figure 3B). The immune cell populations exhibited a strong positive correlation between granulocytes and neutrophils while a low positive correlation existed between NK and NK CD56^bright^ CD16 ^dim/-^There was a strong negative correlation between T cytotoxic and T helper similar to the correlation between NK CD56^dim^ CD16^+^ and NK CD56^dim^ CD16^-^. The NK CD56^dim^ was positively correlated to T helper and negatively correlated to T cytotoxic.

**Figure 3:**
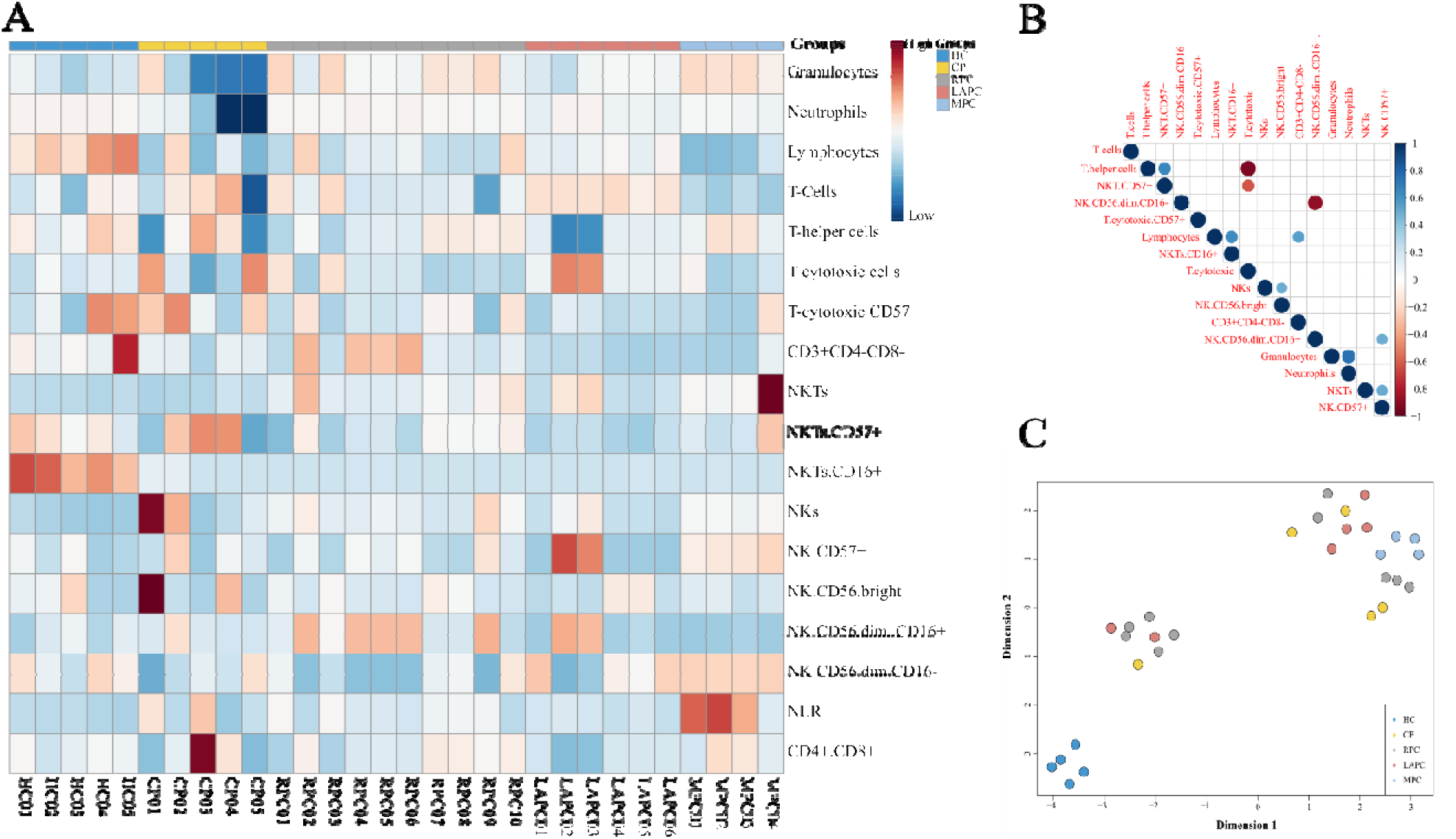
Immune cell Response in PDAC. (A) Heatmap showing the comparison of immune cell markers concentration across the groups from the least concentrated (red) to the most concentrated (blue). The PDAC groups have low concentrations of immune cells compared to the control groups. (B) Correlation matrix of Spearman’s rank correlation coefficients between the immune cell populations. Dark colours indicate strong relationships while weak colours signify weak relationships. (C) Unsupervised clustering of the Immunophenotyping data using KODAMA showed that the controls were distinctively separated from the PDAC groups.

After obtaining the immune matrix, we speculated that these immune cells could distinguish the tumour group from the control group. KODAMA was used to explain the variance-covariance structure of the variable data set through linear combinations of the Immunophenotyping data sets to determine the pattern of separation. MPC, CP, and HC groups were shown to be separate clusters while RPC and LAPC were not distinctively separated (Figure 3C).

### Immune cell ratios in PDAC

Kaplan–Meier survival curves were created using the R library “survival.” The Wald test was used to calculate the *p*-values between survival curves. Prognostic factors for overall survival (OS) were analysed using the Cox proportional hazard regression. Although both CD4/CD8 and neutrophil-lymphocyte ratio (NLR) was not significantly associated with overall survival (OS), the NLR levels increased in RPC, LAPC and MPC when compared with HC (**Figure S2**).

### Dysregulated immune marker genes in PDAC

Gene expression analysis of the PBMCs showed no statistical difference when PDAC groups were compared with the control groups **(Figure S3)**. The CP group had the highest fold change for both *CD56* (2.68± 1.36) and *CD57* (49.22± 63.47). Although there was no statistical correlation or differences observed when comparing the PDAC groups with the control groups, we observed that patients with RPC had the highest foldchange for both *CD4* (11.75 ± 44.31) and *CD3* (30.47 ± 75.01) while LAPC had the highest fold change for *CD8* (3.86 ± 7.35) and *CD16* (51.69 ± 108.9) genes compared to MPC patients.

### Enzyme-Linked Immunosorbent Assay of CD4 and CD8 proteins in plasma

To detect and quantify immunologic reactions in PDAC, the Enzyme-Linked Immunosorbent assay (ELISA) technique was implored on the plasma samples. There were no statistically significant differences observed between the PDAC groups and the controls (**Figure S4**).

### Evaluating the inflammatory status of different PDAC subgroups

The inflammatory status was assessed by analysing the DEPPD levels of PDAC groups and comparing it with the controls. A spectrophotometric assay of ROS showed that there was no significant difference observed when the PDAC groups were compared with CP groups but showed significance when compared with the HC. ROS levels were elevated in RPC (*p*-value = 0.003) and LAPC (*p*-value = 0.008) compared to the HC group (Figure 4A). From a previous study of the analysis of metabolites in PDAC in the same patient cohort, dysregulated metabolites were associated with elevated inflammatory status of the disease (27). Hence, to further understand how and if these metabolites alter the oxidative status of PDAC, DEPPD levels were correlated with the metabolites concentration. Inflammatory markers such as GlycA (rho= 0.45, *p*-value < 0.001) have significantly positive correlation (Figure 4B) while 3-hydroxybutyrate (rho= - 0.45, *p*-value = 0.01) and ascorbate (rho= - 0.56, *p*-value = <0.001) were statistically inversely associated with the DEPPD levels (Figure 4C, **Table S2**). To determine if ROS is a good marker of inflammation, a receiver observing characteristic (ROC) curve was plotted with an area under the curve (AUC) value of 0.91 shown in **Figure S5**.

**Figure 4:**
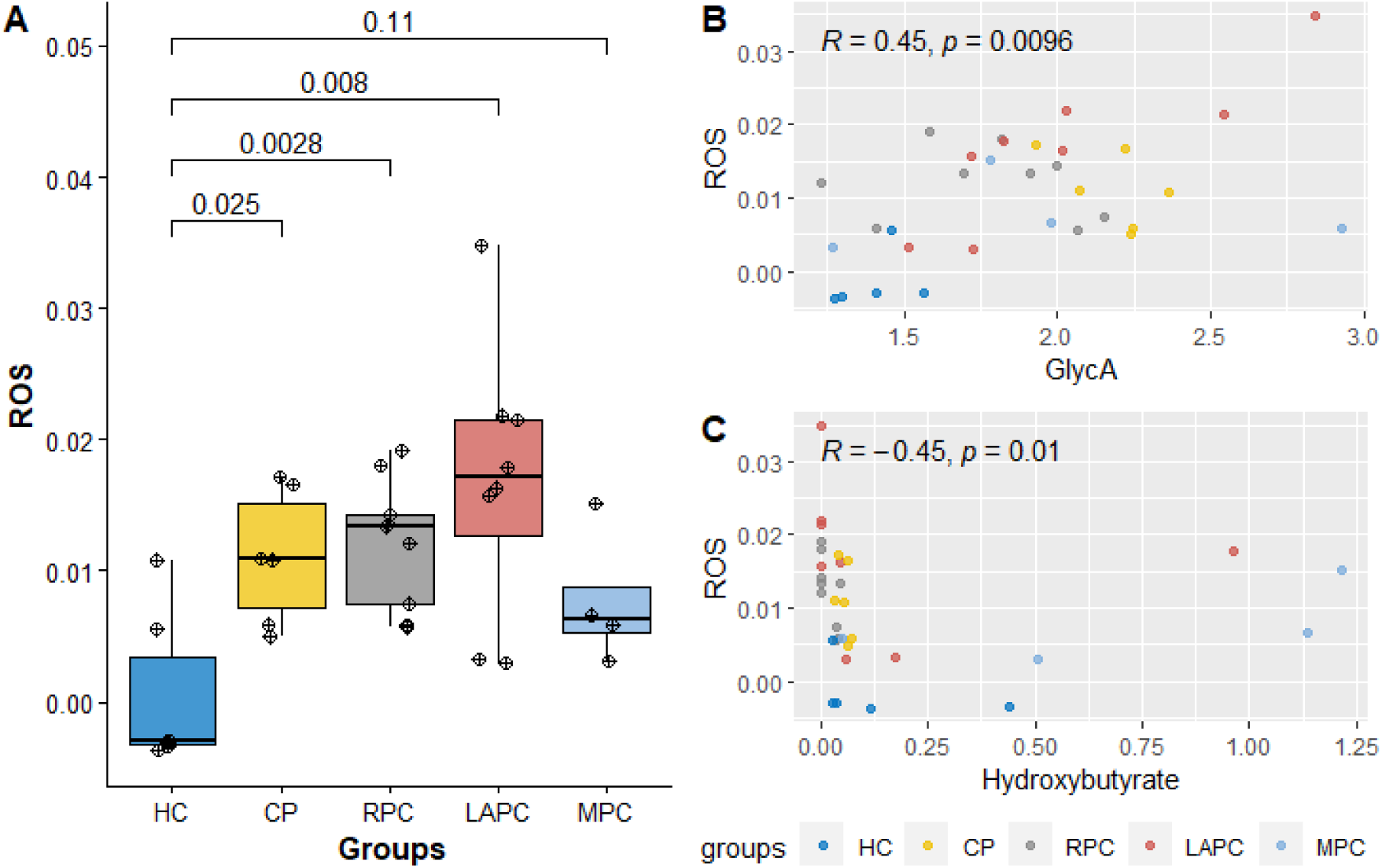
Determination of the inflammatory status of PDAC. **A** Boxplot showing the comparison in DEPPD levels representing ROS activity between the PDAC groups (RPC, LAPC, and MPC) and control groups (HC and CP). A significant change was observed when the RPC and LAPC groups of PDAC were compared with HC groups. **B and C**. Correlation of ROS and inflammatory marker GlycA and 3-hydroxybutyrate respectively. ROS is positively associated with GlycA and negatively correlated with 3-hydroxybutyrate. HC; Healthy controls, CP: Chronic Pancreatitis, RPC: Resectable Pancreatic Ductal Adenocarcinoma, LAPC; Locally Advanced Pancreatic Ductal Adenocarcinoma, MPC; Metastatic Pancreatic Ductal Adenocarcinoma.

## DISCUSSION

PDAC tumour cells exhibit immunosuppressive characteristics (36). Infiltrating immune cells may influence PDAC progression in diverse ways. Hence, the immune status could be essential in predicting the outcome and management of the disease. ROS can either be detrimental or beneficial for immune cell function and response (37). In this study, the expression levels of ROS and immune cell markers PDAC progression were evaluated in comparison to control groups consisting HC and CP.

This study showed that granulocyte levels increased as PDAC progressed from RPC, LAPC to MPC. Recent studies have demonstrated that granulocytes, neutrophils, lymphocytes, and NLR are associated with the overall survival of PDAC patients (9). However, the granulocyte count is an independent predictive factor for PDAC (9). Neutrophils contribute to the majority of the granulocyte population; hence it is reasonable to expect that an elevated granulocyte count in this cohort is a consequence of elevated neutrophil levels with PDAC severity. Neutrophils are the main component of chronic inflammation which promotes tumour initiation and progression (38).

CD16^+^ neutrophils (also known as FcγRIIIb) were observed to increase with the disease severity. Neutrophils can exert both pro and anti-tumoural functions which could depend on the type of tumour and microenvironment (38). Additionally, they exhibit functional plasticity depending on the expression of cell surface markers, cytokines, and ROS. Studies have shown that ROS production is vital in several neutrophil effector functions (39). Neutrophils contribute to the destruction of cancer cells particularly upon treatment with anti-cancer antibodies, however, the existence of immature neutrophils in circulation mediates immunosuppression and subsequent metastasis (39). Neutrophil apoptosis is associated with reduced responsiveness and inhibition of receptors activating effector function (40) and loss of its ability to secrete granule enzymes on deliberate external stimulation (41). CD16^+^ neutrophils uniquely function as an inhibitor of antibody-dependent destruction of cancer cells, thereby identifying it as a potential target for enhancing the therapeutic efficacy of cancer therapeutic antibodies (42).

This study confirms that reduced lymphocyte count is negatively associated with survival rate (9). Pancreatic cancer cells downregulate the immune responses by causing a reduction in total lymphocytes and T helper cells which play a crucial role in immune regulation (43). Except for LAPC, both CD8^+^ T-cells while CD4^+^T-cells were downregulated with tumour severity in this study. Additionally, no statistical alteration was recorded between CD4^+^T-cells and tumour severity for HIV-positive PDAC patients in this cohort. Pancreatic cancer cells escape immunity by secreting cytokines such as IL-10 and TGF-β, immunosuppression as a result of these cytokines affects the immune function by inhibiting the infiltration of CD4^+^ and CD8^+^ T cells in the cancer cells (44). Additionally, DN (CD3^+^ CD4^-^ CD8^-^) T-cells levels, reduced significantly in LAPC and MPC in this cohort. The DN T-cells have been shown to inhibit proliferation and invasion in human pancreatic cancer cells via the Fas/FasL pathway which induces cell apoptosis (45). Furthermore, this study confirms that high NLR suggests a poor prognosis for patients with PDAC and could be used as a novel survival assessment marker (46).

This study showed that NK and NKT cells were significantly elevated in RPC when compared to the controls HC and CP. Furthermore, NKT CD16^+^ cells were significantly reduced across the groups. NK cell activation is accompanied by the secretion of inflammatory cytokines thereby driving inflammation which restricts adaptive immune responses (47). NK cells excluded from PDAC tumours display downregulation of both CD16^+^ and CD57^+^ (48). NK cells fail to survive or proliferate in a hypoxic microenvironment which contributes to the immune escape of NK cells in PDAC patients (49). In the blood of PDAC patients, the downregulation of Natural Killer Group 2D (NKG2D) reduces cytotoxicity, by lowering the levels of IFN-γ with elevated levels of IL-10, an immunoregulatory cytokine (48). Decreased NKG2D levels stimulate impaired killing of the tumour cells by NK and NKT cells (49).

Elevated levels of NK cell markers, CD56^+^ and CD57^+^, correlate with favourable outcomes (50). CD56^+^ drives the maturation of NK cells and is weak in cytotoxicity but strong in the production of anti-tumour cytokines such as IFN-γ and TNF-α (50). However, although not significant the two main CD56^+^ NK subset populations which are; CD56^bright^CD16dim/^-^ and CD56^dim^CD16^+^ subsets were shown to reduce in LAPC and MPC when compared to HC . This might be due to the inhibited immunomodulatory function and cytotoxic capacity of both subsets respectively (17). CD57^+^ NK cells are regarded as a marker of terminal differentiation which is less proliferative but more cytotoxic to tumour cells and could acquire IFN-γ when crosslinked with CD16^+^ (51,52).

### Elevated ROS levels could be vital in PDAC progression

There was an observed elevation of ROS levels as PDAC progressed from RPC to LAPC in this cohort. A mild concentration of ROS promotes cancer progression, while malignant cells induce antioxidant programs such as activation and stabilization of nuclear factor erythroid-derived 2 (Nrf2) to avoid programmed cell death. In response to elevated ROS levels, Kelch-like ECH-associated protein1 (Keap1), which is bound to Nrf2 becomes inactivated (53). Nrf2 then translocates to the nucleus and activates genes that mediate antioxidant programs (53). Hence in PDAC cells, ROS need to be kept at a threshold level that promotes proliferation but prevents senescence and cell death, this could suggest a decrease in ROS levels at the MPC stage. ROS promotes apoptosis and cancer cell survival depending on its concentration and cancer cell type (54). This study confirms that ROS is strongly associated with inflammation because they have closely related pathophysiological activities that are linked (55,56). ROS acts as an inflammatory regulator via the activation of NF-_k_B which promotes the expression of proinflammatory cytokines (57).

There was an observed negative association between ROS and 3-HB and ascorbate. Studies have described the antioxidant role of 3-HB via NADH oxidation which inhibits ROS production (58). Oxidative stress is suppressed via the inhibition of histone deacetylase (HDAC) by 3-HB (59). Ascorbic acid have been shown to inhibit ROS production in cancer cells (60), thereby possessing an antitumour effect.

## CONCLUSION

Effector immune cells could be essential in predicting prognosis in the PDAC cohort. This study showed that increased granulocytes and neutrophil levels, and decreased T-lymphocyte and NK cell levels correlated with poor survival. Evaluating the role of these immune cells as well as their interaction with ROS might be significant in understanding disease progression and in developing novel therapeutic strategies. The small number of recruited patients in each stage is a limitation. However, to our knowledge, this is the first study of its kind in the study population, providing valuable data in this group of patients. Future studies must incorporate a larger patient cohort and further investigate the interplay between the immune cells in inflammation in the tumour microenvironment.

### Authors’ contributions

E.E.N conceptualised the study. P.F., G.C, and E.E.N acquired funding for the project. N.E, P.F, J.OJ and E.E.N designed and collected data. N.E, E.A., S.C., J.D, J.OJ, M.S., Z,N., A.H., P.F., G.C, and E.E.N performed data analysis and interpretation. N.E., E.A., S.C., and E.E.N wrote the initial draft. All authors critically reviewed and approved the final manuscript.

### Funding

Research reported in this publication was supported by the South African Medical Research Council under a Self-Initiated grant, two South African National Research Foundation grants (Grant numbers: 138367 and 121277) and the Cancer Association of South Africa (CANSA). The views and opinions expressed are those of the authors and do not necessarily represent the official views of the funders.

## Data Availability

All data produced in the present study are available upon reasonable request to the authors

## SUPPLEMENTARY TABLES

**Table S1:** 6-colour Flow Cytometry Immunophenotyping Panel.

| Laser Name | Filter | Parameter | Em | lymphocytes | T Cells | NK Cells | B cells, monos & granulocytes | Vol per test (µl) | Optimised volume(µl) | Catalogue #/Clone (2019) |
| --- | --- | --- | --- | --- | --- | --- | --- | --- | --- | --- |
| Red 640 |  |  |  |  |  |  |  |  |  |  |
|  | 730/45 BP | Alexa Fluor 700 | 720 |  | CD4 |  |  | 5 |  | 561030, Clone; RPA-T4 |
| Blue 488 |  |  |  |  |  |  |  |  |  |  |
| A | 780/60 BP | PE-Cy7 | 780 |  |  | CD56 |  | 5 |  | 560916; Clone; B159 |
| C | 660/20 | PE-Cy5 | 670 |  |  | CD16 |  |  |  | 561725, Clone; 3 G8 |
| D | 530/30BP | BB515 | 515 |  | CD57 |  |  | 5 |  | 565285, Clone; NK 1 |
| Violet 405 |  |  |  |  |  |  |  |  |  |  |
| Power 50 | 605/12 | BV605 | 605 |  | CD8 |  |  | 5 |  | 564115, Clone; SK1 |
| UV 355 |  |  |  |  |  |  |  |  |  |  |
|  | 530/30 | BUV496 | 496 | CD3 |  |  |  | 5 |  | 612941, Clone; UCHT1 |

**Table S2:** Percentage population of Immune cell makers in PDAC.

| Samples | Granulocytes (%) | Neutropiles (%) | Lymphocytes (%) | T.cells (%) | T.helper (%) | T.cytotoxic (%) | T.cytotoxic.CD57+ (%) | CD3+CD4-CD8- (%) | NKTs (%) | NKTs.CD57+ (%) | NK (%) | NK.CD57+ (%) | NK.CD56bright (%) | NK.CD56.dim.CD16+ (%) | NK.CD56.dim.CD16- (%) |
| --- | --- | --- | --- | --- | --- | --- | --- | --- | --- | --- | --- | --- | --- | --- | --- |
| HC01 | 59.2 | 98.7 | 25.2 | 56.6 | 63.1 | 34 | 16.3 | 9.79 | 0.89 | 79.8 | 8.41 | 21.7 | 12.5 | 5.29 | 80.3 |
| HC02 | 46.2 | 98.4 | 31.1 | 65.2 | 49.9 | 51.3 | 28.8 | 7.84 | 0.57 | 65.9 | 5.98 | 9.72 | 13.8 | 26.8 | 54.2 |
| HC03 | 34.2 | 98.8 | 26.6 | 45.9 | 55.8 | 38.7 | 6.82 | 9.28 | 0.57 | 50 | 2.37 | 24.4 | 27.8 | 37.2 | 33.8 |
| HC04 | 45.9 | 98.8 | 37.8 | 70.1 | 75.5 | 44.2 | 61.8 | 7.59 | 0.46 | 61.5 | 2.04 | 3 | 0 | 10 | 88 |
| HC05 | 49.1 | 96.2 | 40 | 68.4 | 65.7 | 41.9 | 57.7 | 22.9 | 0.79 | 37.4 | 2.51 | 5.64 | 0 | 30.3 | 67.7 |
| CP01 | 77.6 | 91.1 | 7.01 | 54.8 | 13.9 | 81.4 | 47.7 | 7.82 | 0.24 | 12.5 | 93.9 | 9.06 | 71.1 | 25.4 | 1.44 |
| CP02 | 38.4 | 90.9 | 27.9 | 71.6 | 58.3 | 40.6 | 62.5 | 7.42 | 0.41 | 78.5 | 52.9 | 38.1 | 4.16 | 61 | 31.8 |
| CP03 | 5.55 | 63.5 | 3.85 | 76.3 | 85 | 15.7 | 29 | 3.25 | 0.41 | 100 | 1.32 | 0 | 0 | 25 | 50 |
| CP04 | 2.29 | 10.4 | 16.5 | 85.2 | 67.3 | 29 | 12 | 7.7 | 1.15 | 96.6 | 9.43 | 22 | 34.1 | 22 | 36.6 |
| CP05 | 1.09 | 11.7 | 2.96 | 24.3 | 10.5 | 85 | 46.9 | 7.2 | 0.83 | 0 | 18.7 | 14.1 | 0 | 20.6 | 78.9 |
| RPC01 | 81.8 | 97.6 | 9.63 | 74.3 | 35.3 | 61.7 | 14.9 | 4.07 | 2.4 | 8.07 | 24 | 3.24 | 13.9 | 29.9 | 54.4 |
| RPC02 | 51.5 | 96.8 | 29.9 | 62.3 | 57 | 28.7 | 39.8 | 15.1 | 6.06 | 59.5 | 32.6 | 21.4 | 5.98 | 82.8 | 10.6 |
| RPC03 | 81.1 | 97.4 | 9.76 | 76.5 | 35.2 | 62.5 | 11.8 | 4.71 | 2.12 | 18.1 | 19.4 | 5.23 | 17.6 | 46.1 | 34.2 |
| RPC04 | 63.3 | 97.9 | 16.4 | 57.6 | 47.3 | 39.6 | 20.7 | 14.1 | 1.99 | 32.8 | 20.1 | 23.4 | 8.31 | 80.4 | 10.6 |
| RPC05 | 62 | 97.7 | 16 | 58.1 | 49 | 39 | 17.7 | 14 | 2.01 | 34.2 | 23.4 | 18.6 | 7.37 | 80.9 | 10.8 |
| RPC06 | 60.3 | 98.2 | 18.3 | 59.7 | 47.2 | 39.6 | 18.8 | 15.3 | 2.18 | 21.4 | 21.7 | 22.7 | 6.04 | 81.2 | 11.5 |
| RPC07 | 75.5 | 97.8 | 11.4 | 59.7 | 64 | 29.6 | 34.2 | 7.07 | 3.11 | 52.9 | 14.8 | 6.21 | 20.6 | 24 | 53.1 |
| RPC08 | 71.7 | 97.3 | 10.7 | 63.1 | 63.4 | 30.6 | 33.9 | 6.45 | 2.12 | 51.5 | 15.9 | 5.23 | 16.2 | 24.8 | 57.7 |
| RPC09 | 79.5 | 95.8 | 10.5 | 39.9 | 62.4 | 34.2 | 5.72 | 5.16 | 3.35 | 50.8 | 42.2 | 31.6 | 6.57 | 84.8 | 5.85 |
| RPC10 | 57.3 | 96.8 | 29.2 | 64.5 | 63.9 | 30.6 | 39.8 | 7.37 | 4.33 | 57.5 | 27.6 | 14 | 6.81 | 14.7 | 74.1 |
| LAPC01 | 50.7 | 88.9 | 16.7 | 75.3 | 49 | 48 | 10.3 | 4.5 | 0.8 | 21.1 | 11.1 | 0.98 | 1.97 | 2.46 | 94.1 |
| LAPC02 | 41.4 | 98.7 | 16.2 | 75 | 9.07 | 87.4 | 25.1 | 4.21 | 4.21 | 27.8 | 25.2 | 65.9 | 4.88 | 87.4 | 5.76 |
| LAPC03 | 61.1 | 98.1 | 18.3 | 74.5 | 11.9 | 84.3 | 20.2 | 4.43 | 5.09 | 28.8 | 33.7 | 54.5 | 3.07 | 80 | 15.7 |
| LAPC04 | 62.1 | 91.8 | 21.4 | 76.5 | 50.9 | 46.5 | 14.8 | 3.26 | 1.02 | 20.4 | 11.7 | 1.2 | 21.6 | 4.8 | 67 |
| LAPC05 | 62.8 | 91.4 | 16.8 | 70.7 | 51 | 46.4 | 16.3 | 3.21 | 0.86 | 16.1 | 12 | 1 | 21.6 | 11.8 | 63.5 |
| LAPC06 | 61 | 90.4 | 21.7 | 74.3 | 50.9 | 46.7 | 15.6 | 3.34 | 1.21 | 28.5 | 8.45 | 0.69 | 4.63 | 3.24 | 90.7 |
| MPC01 | 79.2 | 94.7 | 3.94 | 53.3 | 53.3 | 53.3 | 15.1 | 3.4 | 3.19 | 38.7 | 27.5 | 28.2 | 10 | 0 | 89.5 |
| MPC02 | 77.1 | 94.5 | 3.64 | 49.5 | 68.8 | 28.3 | 10.2 | 3.06 | 2.94 | 28.6 | 26.6 | 30.3 | 10.4 | 3.1 | 86.5 |
| MPC03 | 78.2 | 94.4 | 5.02 | 54.1 | 67 | 30.4 | 9.81 | 3.06 | 3.27 | 32.5 | 23.5 | 32.3 | 4.53 | 7 | 86.6 |
| MPC04 | 67.4 | 91.5 | 13.5 | 49.7 | 52.5 | 38.7 | 41.1 | 7.83 | 12 | 79.2 | 27.9 | 37.1 | 6.97 | 2.06 | 89.9 |

**Table S3:** DEPPD Levels correlation with Metabolites concentration in PDAC.

| **Feature** | **Rho** | **p-value** | **FDR** |
| --- | --- | --- | --- |
| Formate | 0.15 | 0.418 | 0.889 |
| Unknown signal at 8.12 ppm | -0.08 | 0.652 | 0.922 |
| Unknown signal at 8.07 ppm | 0.09 | 0.618 | 0.922 |
| Phenylalanine | 0.06 | 0.724 | 0.922 |
| Tyrosine | -0.24 | 0.187 | 0.889 |
| Unknown signal at 7.14 ppm | -0.02 | 0.926 | 0.945 |
| Histidine | -0.27 | 0.134 | 0.889 |
| Glucose | 0.04 | 0.831 | 0.941 |
| Mannose | 0.15 | 0.415 | 0.889 |
| Unknown signal at 5.15 ppm | -0.11 | 0.553 | 0.922 |
| Unknown signal at 5.09 ppm | 0.01 | 0.970 | 0.971 |
| Unknown signal at 5.01 ppm | -0.03 | 0.891 | 0.941 |
| Ascorbate | -0.56 | <0.001 | 0.042 |
| Threonine | -0.19 | 0.287 | 0.889 |
| Lactate | 0.02 | 0.896 | 0.941 |
| Creatinine | -0.19 | 0.296 | 0.889 |
| Creatine | -0.18 | 0.318 | 0.889 |
| Glycine | 0.08 | 0.682 | 0.922 |
| Methanol | -0.09 | 0.612 | 0.922 |
| Unknown signal at 2.55 ppm | -0.07 | 0.711 | 0.922 |
| Citrate | -0.24 | 0.190 | 0.889 |
| Glutamine | -0.3 | 0.092 | 0.889 |
| Pyruvate | -0.1 | 0.585 | 0.922 |
| Glutamate | 0.1 | 0.579 | 0.922 |
| Acetoacetate | 0.03 | 0.888 | 0.941 |
| Acetate | -0.23 | 0.215 | 0.889 |
| Alanine | -0.16 | 0.373 | 0.889 |
| Unknown signal at 1.45 ppm | 0.28 | 0.119 | 0.889 |
| Unknown signal at 1.43 ppm | -0.02 | 0.904 | 0.941 |
| 3-Hydroxybutyrate | -0.06 | 0.733 | 0.922 |
| Ethanol | 0.03 | 0.892 | 0.941 |
| Unknown signal at 1.16 ppm | 0.06 | 0.753 | 0.922 |
| Unknown signal at 1.14 ppm | 0.13 | 0.490 | 0.922 |
| Unknown signal at 1.11 ppm | 0.16 | 0.388 | 0.889 |
| Unknown signal at 1.06 ppm | -0.27 | 0.139 | 0.889 |
| Valine | -0.22 | 0.230 | 0.889 |
| Isoleucine | 0.06 | 0.727 | 0.922 |
| Leucine | -0.05 | 0.792 | 0.940 |
| 2-Hydroxybutyrate | -0.14 | 0.445 | 0.907 |
| Protein NH | -0.19 | 0.287 | 0.889 |
| Unsaturated lipid -CH=CH- | -0.15 | 0.418 | 0.889 |
| Lipid alpha-CH2 | 0.06 | 0.741 | 0.922 |
| Cholesterol | -0.21 | 0.257 | 0.889 |
| Lipid =CH-CH2-CH= | -0.11 | 0.542 | 0.922 |
| Glycerol phospholipid | 0.06 | 0.742 | 0.922 |
| Phospholipid | 0.28 | 0.127 | 0.889 |
| Lipid beta-CH2 | -0.23 | 0.197 | 0.889 |
| Lipid CH2 | -0.06 | 0.759 | 0.922 |
| Lipid CH3 | -0.17 | 0.342 | 0.889 |
| GlycB | 0.48 | 0.005 | 0.091 |
| GlycA | 0.51 | 0.003 | 0.070 |

## Supplementary Figures

**Figure S1:**
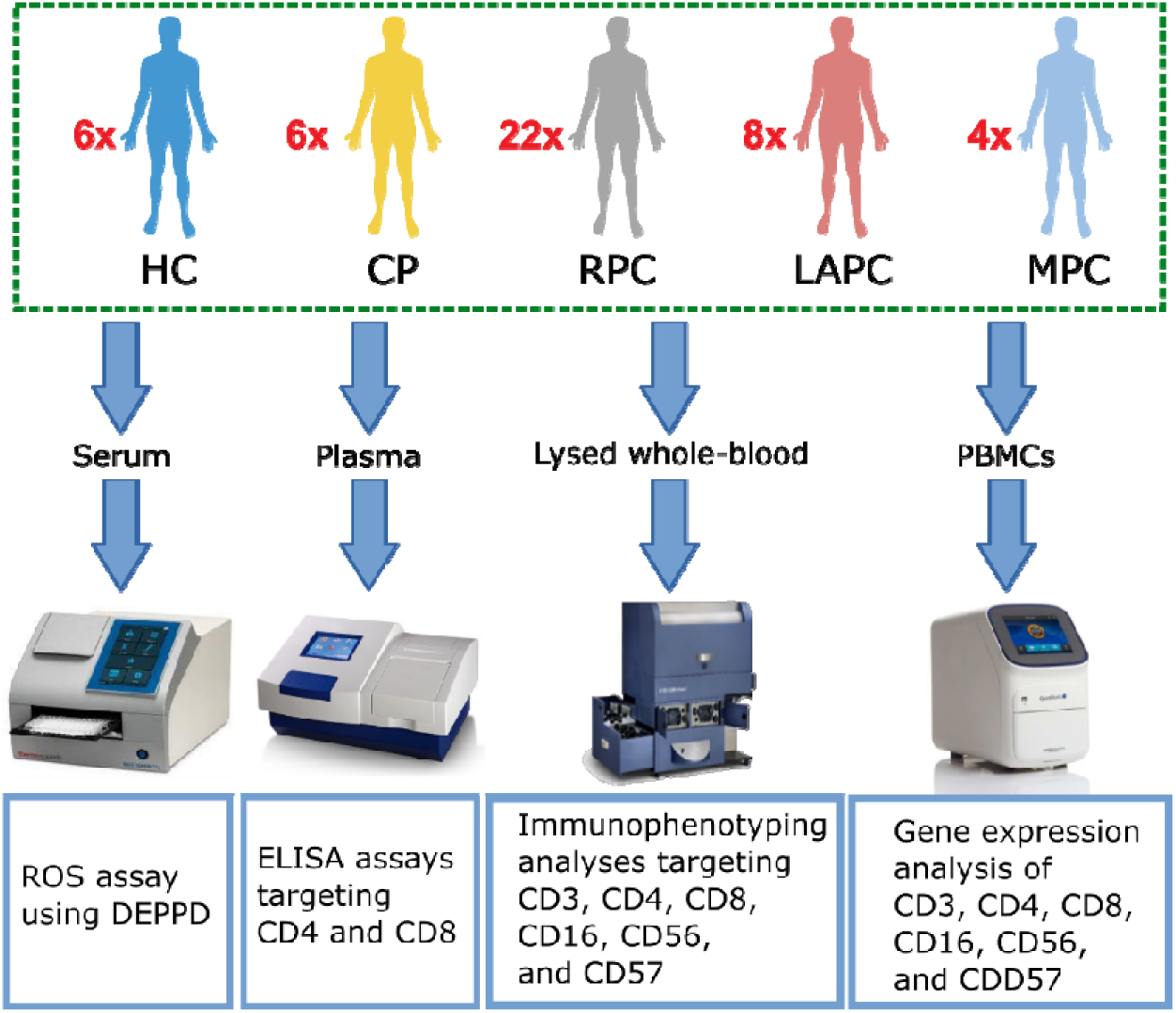
Overview of Sample Size and Processing. After Ethics approval and informed consent blood samples collected were processed into serum, plasma, and PBMCs for ROS, Elisa, and Real-Time Polymerase Chain Reaction Analyses respectively. Whole blood was lysed to fix the white blood cells for Immunophenotyping assays. HC; Healthy controls, CP: Chronic Pancreatitis, RPC: Resectable Pancreatic Ductal Adenocarcinoma, LAPC; Locally Advanced Pancreatic Ductal Adenocarcinoma, MPC; Metastatic Pancreatic Ductal Adenocarcinoma.

**Figure S2:**
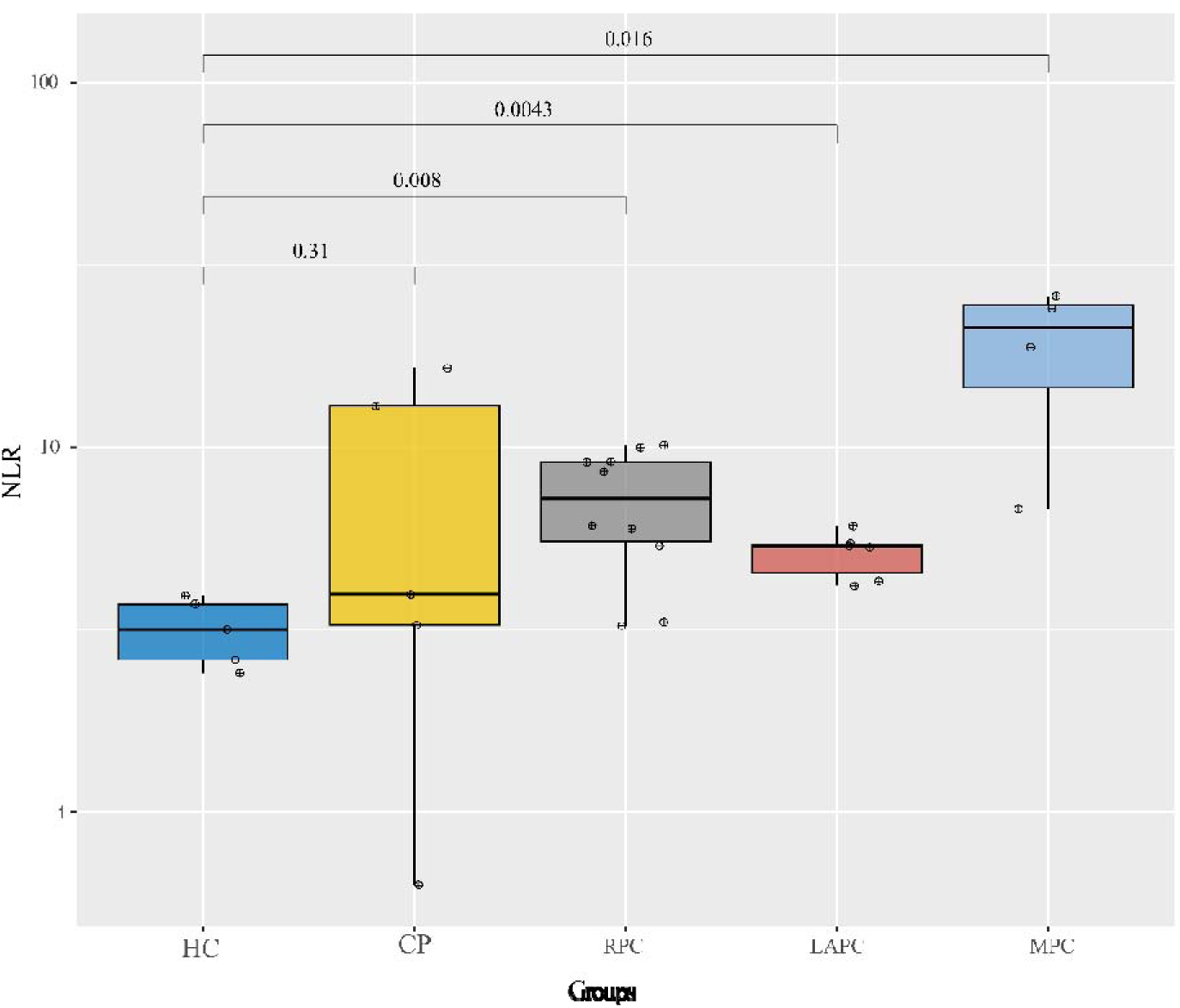
Neutrophil/Lymphocyte Ratio (NLR) across PDAC groups. Elevated levels of NLR were observed as PDAC severity increased and this might signify poor prognosis and could be used as a marker for survival evaluation.

**Figure S3:**
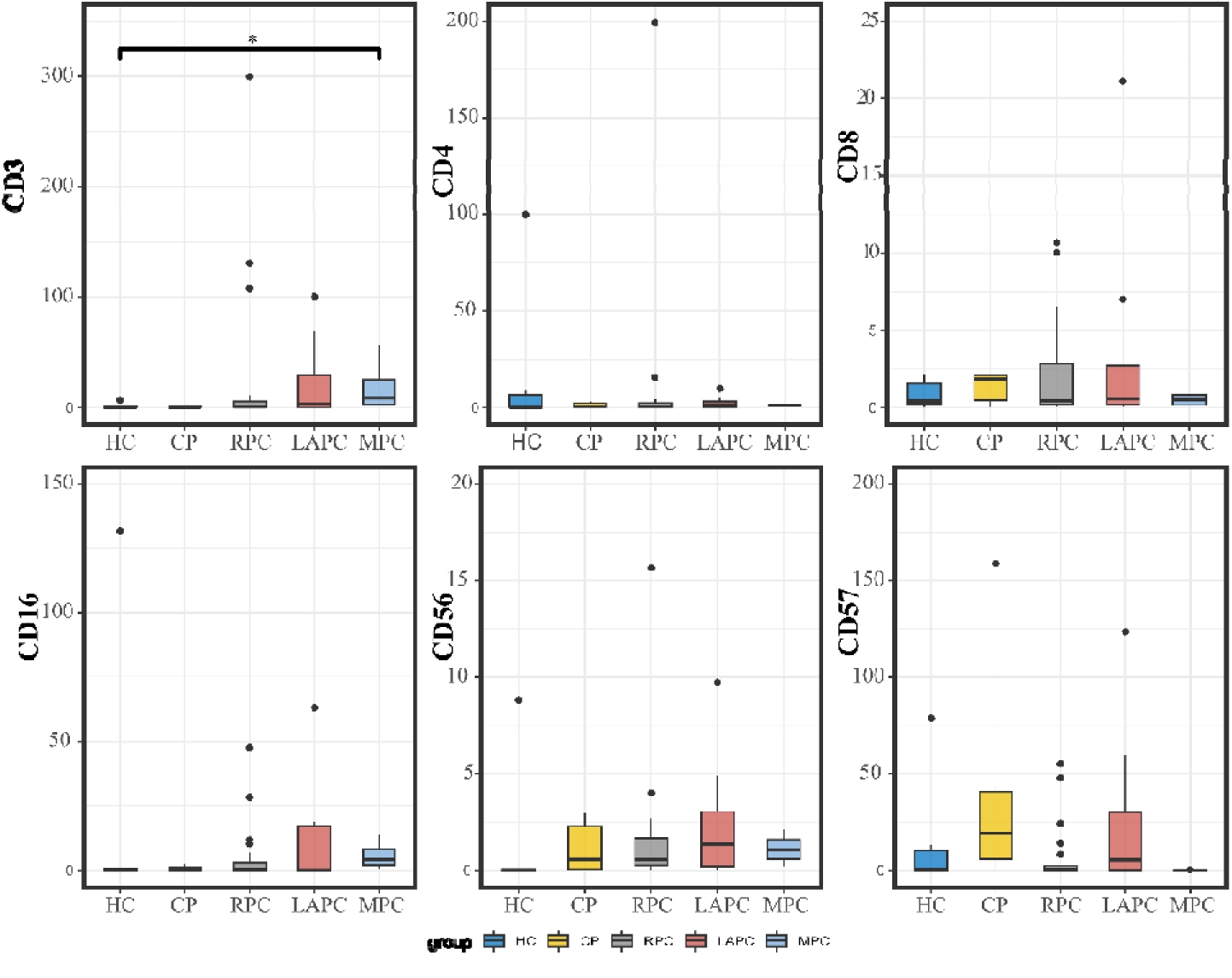
Graphical representation of the immune marker by gene expression. Although there was no statistical correlation or differences observed when comparing the PDAC groups with the control groups for all the markers except for CD3, there were alterations observed across the groups. RPC: Resectable Pancreatic Adenocarcinoma; LAPC: Locally Advanced Pancreatic Adenocarcinoma; MPC: Metastatic Pancreatic Adenocarcinoma., HC: Healthy Controls *p < 0.05, **p < 0.01, ***p < 0.001, n.s., not significant

**Figure S4:**
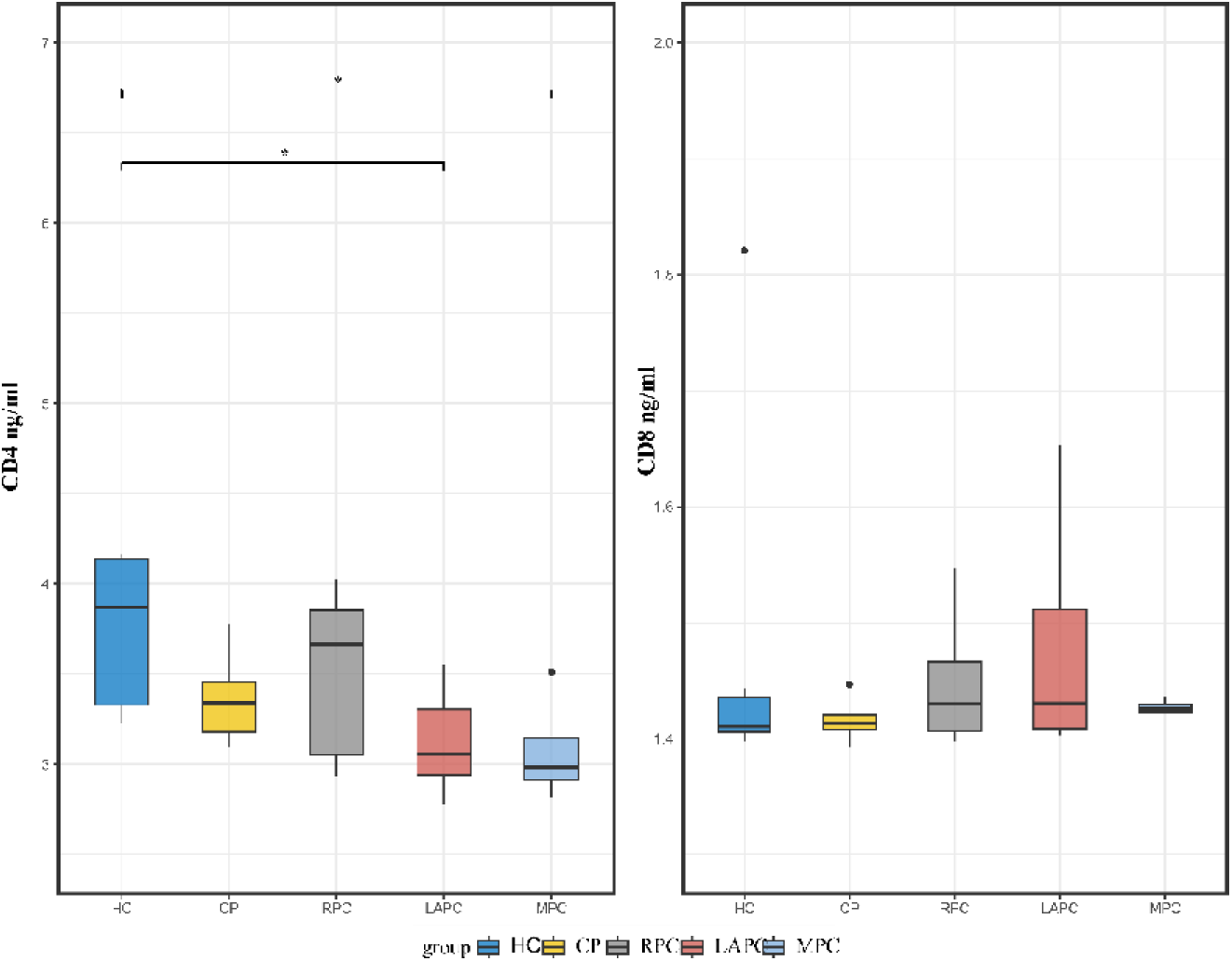
Boxplots comparing the immune cell markers in PDAC plasma samples. Enzyme**-**Linked Immunosorbent Assays (ELISA) technique was implored on the plasma samples. There were no statistically significant differences observed between the PDAC groups and the controls. *p < 0.05, **p < 0.01, ***p < 0.001, n.s., not significant

**Figure S5:**
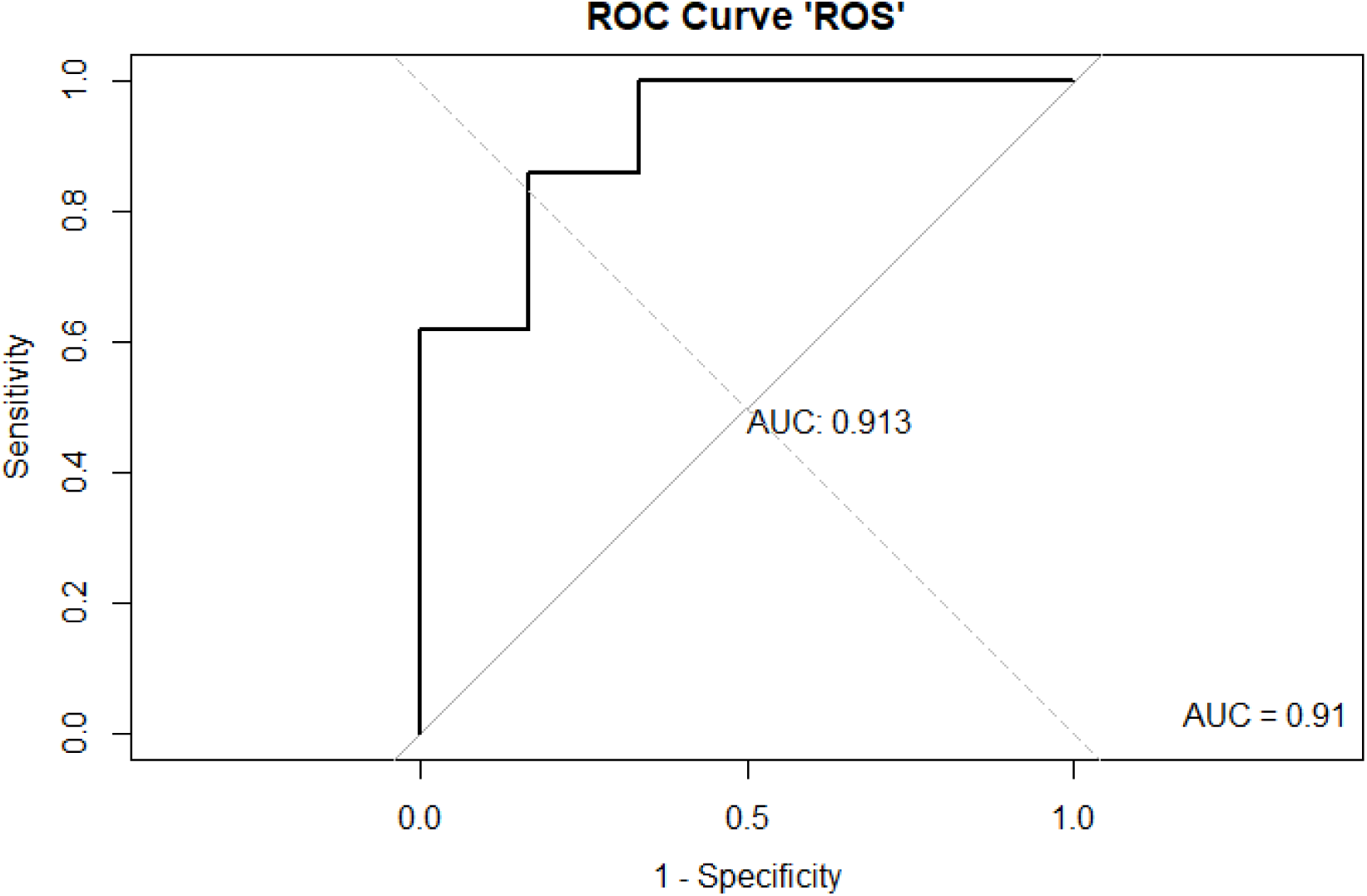
ROC analysis of the ROS plot. AUC of the plot was 0.91 which indicates an outstanding plot. Hence this confirms that ROS is a good marker of inflammation

## REFERENCES

1. Siegel RL, Miller KD, Wagle NS, Jemal A. Cancer statistics, 2023. CA: A Cancer Journal for Clinicians. 2023 Jan 1;73(1):17–48.

2. Rahib L, Smith BD, Aizenberg R, Rosenzweig AB, Fleshman JM, Matrisian LM. Projecting Cancer Incidence and Deaths to 2030: The Unexpected Burden of Thyroid, Liver, and Pancreas Cancers in the United States. Cancer Res. 2014 Jun 1;74(11):2913.

3. Hingorani SR. Epithelial and stromal co-evolution and complicity in pancreatic cancer. Nature Reviews Cancer. 2023 Feb 1;23(2):57–77.

4. Zhu YH, Zheng JH, Jia QY, Duan ZH, Yao HF, Yang J, et al. Immunosuppression, immune escape, and immunotherapy in pancreatic cancer: focused on the tumor microenvironment. Cellular Oncology. 2023 Feb 1;46(1):17–48.

5. Neoptolemos JP, Kleeff J, Michl P, Costello E, Greenhalf W, Palmer DH. Therapeutic developments in pancreatic cancer: current and future perspectives. Nat Rev Gastroenterol Hepatol. 2018 Jun;15(6):333–48.

6. Wei K, Hackert T. Surgical Treatment of Pancreatic Ductal Adenocarcinoma. Cancers. 2021;13(8).

7. Elebo N, Fru P, Omoshoro-Jones J, Patrick Candy G, Nweke EE. Role of different immune cells and metabolic pathways in modulating the immune response in pancreatic cancer (Review). Mol Med Rep. 2020 Dec;22(6):4981–91.

8. Nsingwane Z, Candy G, Devar J, Omoshoro-Jones J, Smith M, Nweke E. Immunotherapeutic strategies in pancreatic ductal adenocarcinoma (PDAC): current perspectives and future prospects. Molecular Biology Reports. 2020 Aug 1;47(8):6269–80.

9. Feng L, Gu S, Wang P, Chen H, Chen Z, Meng Z, et al. White Blood Cell and Granulocyte Counts Are Independent Predictive Factors for Prognosis of Advanced Pancreatic Caner. M’Koma A, editor. Gastroenterology Research and Practice. 2018 May 8;2018:8096234.

10. Shin K, Jung EK, Park SJ, Jeong S, Kim IH, Lee MA. Neutrophil-to-lymphocyte ratio and carbohydrate antigen 19-9 as prognostic markers for advanced pancreatic cancer patients receiving first-line chemotherapy. World J Gastrointest Oncol. 2021 Aug;13(8):915–28.

11. Chang JH, Jiang Y, Pillarisetty VG. Role of immune cells in pancreatic cancer from bench to clinical application: An updated review. Medicine (Baltimore). 2016 Dec;95(49):e5541–e5541.

12. Xiang H, Yang R, Tu J, Xi Y, Yang S, Lv L, et al. Metabolic reprogramming of immune cells in pancreatic cancer progression. Biomedicine & Pharmacotherapy. 2023 Jan 1;157:113992.

13. Garrido F, Perea F, Bernal M, Sánchez-Palencia A, Aptsiauri N, Ruiz-Cabello F. The Escape of Cancer from T Cell-Mediated Immune Surveillance: HLA Class I Loss and Tumor Tissue Architecture. Vaccines. 2017;5(1).

14. Yang F, Feng C, Zhang X, Lu J, Zhao Y. The Diverse Biological Functions of Neutrophils, Beyond the Defense Against Infections. Inflammation. 2017 Feb 1;40(1):311–23.

15. Paul S, Lal G. The Molecular Mechanism of Natural Killer Cells Function and Its Importance in Cancer Immunotherapy. Front Immunol. 2017 Sep 13;8:1124–1124.

16. Abel AM, Yang C, Thakar MS, Malarkannan S. Natural Killer Cells: Development, Maturation, and Clinical Utilization. Front Immunol. 2018;9:1869.

17. Granzin M, Wagner J, Köhl U, Cerwenka A, Huppert V, Ullrich E. Shaping of Natural Killer Cell Antitumor Activity by *Ex Vivo* Cultivation. Front Immunol. 2017;8:458.

18. Wörmann SM, Diakopoulos KN, Lesina M, Algül H. The immune network in pancreatic cancer development and progression. Oncogene. 2013 Jul 15;33:2956.

19. Fru PN, Nweke EE, Augustine TN. Harnessing the Tumor Microenvironment for Cancer Immunotherapy. In: Rezaei N, editor. Handbook of Cancer and Immunology [Internet]. Cham: Springer International Publishing; 2022. p. 1–25. Available from: 10.1007/978-3-030-80962-1_183-1

20. Schlanger D, Popa C, Palca S, Seicean A, Al Hajjar N. The role of systemic immuno-inflammatory factors in resectable pancreatic adenocarcinoma: a cohort retrospective study. World Journal of Surgical Oncology. 2022 May 6;20(1):144.

21. Oberkampf M, Guillerey C, Mouriès J, Rosenbaum P, Fayolle C, Bobard A, et al. Mitochondrial reactive oxygen species regulate the induction of CD8(+) T cells by plasmacytoid dendritic cells. Nat Commun. 2018 Jun 8;9(1):2241–2241.

22. Cheung EC, DeNicola GM, Nixon C, Blyth K, Labuschagne CF, Tuveson DA, et al. Dynamic ROS Control by TIGAR Regulates the Initiation and Progression of Pancreatic Cancer. Cancer Cell. 2020 Feb 10;37(2):168–182.e4.

23. Park HJ, Choi YJ, Lee JH, Nam MJ. Naringenin causes ASK1-induced apoptosis via reactive oxygen species in human pancreatic cancer cells. Food and Chemical Toxicology. 2017 Jan 1;99:1–8.

24. Martinez-Useros J, Li W, Cabeza-Morales M, Garcia-Foncillas J. Oxidative Stress: A New Target for Pancreatic Cancer Prognosis and Treatment. J Clin Med. 2017 Mar 9;6(3):29.

25. Matilla AJ. Cellular oxidative stress in programmed cell death: focusing on chloroplastic 1O2 and mitochondrial cytochrome-c release. Journal of Plant Research. 2021 Mar 1;134(2):179–94.

26. Kiely M, Lord B, Ambs S. Immune response and inflammation in cancer health disparities. Trends in Cancer. 2022 Apr 1;8(4):316–27.

27. Elebo N, Omoshoro-Jones J, Fru PN, Devar J, De Wet van Zyl C, Vorster BC, et al. Serum Metabolomic and Lipoprotein Profiling of Pancreatic Ductal Adenocarcinoma Patients of African Ancestry. Metabolites. 2021;11(10).

28. Yao S, Hong CC, Ruiz-Narváez EA, Evans SS, Zhu Q, Schaefer BA, et al. Genetic ancestry and population differences in levels of inflammatory cytokines in women: Role for evolutionary selection and environmental factors. PLoS Genet. 2018 Jun;14(6):e1007368.

29. Kleiveland CR. Peripheral Blood Mononuclear Cells. In: Verhoeckx K, Cotter P, López-Expósito I, Kleiveland C, Lea T, Mackie A, et al., editors. The Impact of Food Bioactives on Health: in vitro and ex vivo models [Internet]. Cham: Springer International Publishing; 2015. p. 161–7. Available from: 10.1007/978-3-319-16104-4_15

30. Nalisa M, Nweke EE, Smith MD, Omoshoro-Jones J, Devar JW, Metzger R, et al. Chemokine receptor 8 expression may be linked to disease severity and elevated interleukin 6 secretion in acute pancreatitis. World J Gastrointest Pathophysiol. 2021 Nov;12(6):115–33.

31. Mohelnikova-Duchonova B, Oliverius M, Honsova E, Soucek P. Evaluation of reference genes and normalization strategy for quantitative real-time PCR in human pancreatic carcinoma. Dis Markers. 2012;32(3):203–10.

32. Bustin SA, Benes V, Garson JA, Hellemans J, Huggett J, Kubista M, et al. The MIQE Guidelines: Minimum Information for Publication of Quantitative Real-Time PCR Experiments. Clinical Chemistry. 2009 Apr 1;55(4):611–22.

33. Hayashi I, Morishita Y, Imai K, Nakamura M, Nakachi K, Hayashi T. High-throughput spectrophotometric assay of reactive oxygen species in serum. Mutation Research/Genetic Toxicology and Environmental Mutagenesis. 2007 Jul 10;631(1):55–61.

34. Verde V, Fogliano V, Ritieni A, Maiani G, Morisco F, Caporaso N. Use of N, N -dimethyl-p - phenylenediamine to Evaluate the Oxidative Status of Human Plasma. Free Radical Research. 2002 Jan 1;36(8):869–73.

35. Herold NC, Mitra P. Immunophenotyping [Internet]. StatPearls Publishing, Treasure Island (FL); 2022. Available from: http://europepmc.org/abstract/MED/32644353

36. Foucher ED, Ghigo C, Chouaib S, Galon J, Iovanna J, Olive D. Pancreatic Ductal Adenocarcinoma: A Strong Imbalance of Good and Bad Immunological Cops in the Tumor Microenvironment. Front Immunol. 2018 May 14;9:1044–1044.

37. Kotsafti A, Scarpa M, Castagliuolo I, Scarpa M. Reactive Oxygen Species and Antitumor Immunity—From Surveillance to Evasion. Cancers. 2020;12(7).

38. Galdiero MR, Varricchi G, Loffredo S, Mantovani A, Marone G. Roles of neutrophils in cancer growth and progression. Journal of Leukocyte Biology. 2018 Mar 1;103(3):457–64.

39. Mackey JBG, Coffelt SB, Carlin LM. Neutrophil Maturity in Cancer. Front Immunol. 2019 Aug 14;10:1912–1912.

40. Hart SP, Ross JA, Ross K, Haslett C, Dransfield I. Molecular characterization of the surface of apoptotic neutrophils: Implications for functional downregulation and recognition by phagocytes. Cell Death & Differentiation. 2000 May 1;7(5):493–503.

41. Haslett C, Savill JS, Whyte MKB, Stern M, Dransfield I, Meagher LC, et al. Granulocyte apoptosis and the control of inflammation. Philosophical Transactions of the Royal Society of London Series B: Biological Sciences. 1994 Aug 30;345(1313):327–33.

42. Treffers LW, van Houdt M, Bruggeman CW, Heineke MH, Zhao XW, van der Heijden J, et al. FcγRIIIb Restricts Antibody-Dependent Destruction of Cancer Cells by Human Neutrophils. Front Immunol. 2019 Jan 30;9:3124–3124.

43. Fogar P, Sperti C, Basso D, Sanzari MC, Greco E, Davoli C, et al. Decreased Total Lymphocyte Counts in Pancreatic Cancer: An Index of Adverse Outcome. Pancreas [Internet]. 2006;32(1). Available from: https://journals.lww.com/pancreasjournal/fulltext/2006/01000/decreased_total_lymphocyte_counts_in_pancreatic.4.aspx

44. Fukunaga A, Miyamoto M, Cho Y, Murakami S, Kawarada Y, Oshikiri T, et al. CD8+tumor-infiltrating lymphocytes together with CD4+tumor-infiltrating lymphocytes and dendritic cells improve the prognosis of patients with pancreatic adenocarcinoma. Pancreas. 2004;28(1):e26–31.

45. YIN LU, PIBO HU, HAIBO ZHOU, ZHIJIAN YANG, YU SUN, ROBERT M. HOFFMAN, et al. Double-negative T Cells Inhibit Proliferation and Invasion of Human Pancreatic Cancer Cells in Co-culture. Anticancer Res. 2019 Nov 1;39(11):5911.

46. Yang JJ, Hu ZG, Shi WX, Deng T, He SQ, Yuan SG. Prognostic significance of neutrophil to lymphocyte ratio in pancreatic cancer: a meta-analysis. World J Gastroenterol. 2015 Mar 7;21(9):2807–15.

47. Zitti B, Bryceson YT. Natural killer cells in inflammation and autoimmunity. Cytokine & Growth Factor Reviews. 2018 Aug 1;42:37–46.

48. Marcon F, Zuo J, Pearce H, Nicol S, Margielewska-Davies S, Farhat M, et al. NK cells in pancreatic cancer demonstrate impaired cytotoxicity and a regulatory IL-10 phenotype. null. 2020 Jan 1;9(1):1845424.

49. Lim SA, Kim J, Jeon S, Shin MH, Kwon J, Kim TJ, et al. Defective Localization With Impaired Tumor Cytotoxicity Contributes to the Immune Escape of NK Cells in Pancreatic Cancer Patients. Front Immunol. 2019;10:496.

50. Zhang S, Liu W, Hu B, Wang P, Lv X, Chen S, et al. Prognostic Significance of Tumor-Infiltrating Natural Killer Cells in Solid Tumors: A Systematic Review and Meta-Analysis. Front Immunol. 2020 Jul 2;11:1242–1242.

51. Kared H, Martelli S, Tan SW, Simoni Y, Chong ML, Yap SH, et al. Adaptive NKG2C(+)CD57(+) Natural Killer Cell and Tim-3 Expression During Viral Infections. Front Immunol. 2018 Apr 20;9:686–686.

52. Kared H, Martelli S, Ng TP, Pender SLF, Larbi A. CD57 in human natural killer cells and T-lymphocytes. Cancer Immunol Immunother. 2016 Apr;65(4):441–52.

53. Durand N, Storz P. Targeting reactive oxygen species in development and progression of pancreatic cancer. null. 2017 Jan 2;17(1):19–31.

54. Aggarwal V, Tuli HS, Varol A, Thakral F, Yerer MB, Sak K, et al. Role of Reactive Oxygen Species in Cancer Progression: Molecular Mechanisms and Recent Advancements. Biomolecules. 2019;9(11).

55. Menzel A, Samouda H, Dohet F, Loap S, Ellulu MS, Bohn T. Common and Novel Markers for Measuring Inflammation and Oxidative Stress Ex Vivo in Research and Clinical Practice—Which to Use Regarding Disease Outcomes? Antioxidants. 2021;10(3).

56. Biswas SK. Metabolic Reprogramming of Immune Cells in Cancer Progression. Immunity. 2015 Sep 15;43(3):435–49.

57. Forrester SJ, Kikuchi DS, Hernandes MS, Xu Q, Griendling KK. Reactive Oxygen Species in Metabolic and Inflammatory Signaling. Circ Res. 2018 Mar;122(6):877–902.

58. Rojas-Morales P, Pedraza-Chaverri J, Tapia E. Ketone bodies, stress response, and redox homeostasis. Redox Biology. 2020 Jan 1;29:101395.

59. Shimazu T, Hirschey MD, Newman J, He W, Shirakawa K, Le Moan N, et al. Suppression of oxidative stress by β-hydroxybutyrate, an endogenous histone deacetylase inhibitor. Science. 2013 Jan;339(6116):211–4.

60. Fukumura H, Sato M, Kezuka K, Sato I, Feng X, Okumura S, et al. Effect of ascorbic acid on reactive oxygen species production in chemotherapy and hyperthermia in prostate cancer cells. The Journal of Physiological Sciences. 2012 May 1;62(3):251–7.

